# Knowledge, attitude, and practices towards COVID-19 among the Rohingya refugees in Cox’s Bazar, Bangladesh

**DOI:** 10.1101/2021.07.23.21260991

**Authors:** Md. Fahad Jubayer, Md. Tariqul Islam Limon, Md. Masud Rana, Md. Shahidullah Kayshar, Md. Shoaib Arifin, AHM Musleh Uddin, Md. Anisur Rahman Mazumder

## Abstract

The Rohingya refugee population in Bangladesh has become more vulnerable to COVID-19 because of their living and environmental conditions. The current study represents an assessment of the Rohingya people’s COVID-19-related knowledge, attitude, and practices (KAP) at eight refugee camps in Cox’s Bazar. This cross-sectional study was completed with a total of 400 responses between July and September of 2020. A questionnaire was created to assess demographic characteristics (5 items), knowledge (10 items), attitude (5 items), practices (5 items), and information sources (1 item). Aside from the total KAP scores, the scores are also presented based on demographic variables. The KAP of the respondents were not satisfactory, with scores of 5.8 ±1.8, 2.2 ± 1.0, and 0.9 ± 0.7, respectively. We found significant differences only in the knowledge scores based on education and gender. In conclusion, this study emphasizes the importance of COVID-19 training that focuses on behavioral changes for the Rohingya people in Bangladesh.

## Introduction

Coronavirus disease 2019 (COVID-19) has become a worldwide public health concern since its discovery in China in December 2019. It is caused by a novel coronavirus known as severe acute respiratory syndrome coronavirus 2 (SARS-COV-2) (Cucinotta and Vanelli 2020; Lake 2020). Primarily, COVID 19 transmitted by respiratory droplets in person to person showing fever, fatigue, breathing distress, and respiratory complications (Huang et al. 2020). Bangladesh, like the rest of the world, is witnessing the annihilation of this lethal virus. Underprivileged people suffer the most in situations like this. Displaced refugee populations may be at the top of the list of those who are disadvantaged. The COVID-19 pandemic has been declared a global health emergency by WHO, and the virus poses a significant risk to Rohingya refugees in Bangladesh.

A large number of Rohingya refugees are currently residing in Cox’s Bazar, Bangladesh, with inadequate access to water and sanitation (Jubayer et al., 2021). A lack of health and hygiene infrastructure, as well as a health information system, may all contribute to the spread of this disease within the camps. Fear and misinformation about the COVID-19 may exacerbate the situation in the camps, given the current telecommunications blockage, limited Internet access within the camp area, and the Rohingya people’s illiteracy (Limon et al., 2020). The entire refugee accommodation area in Cox’s Bazar has characteristics that make it a likely epicentre of infection spread.

People must follow the World Health Organization’s control measures for the successful prevention and control of this coronavirus disease (WHO, 2020). The effectiveness of precautionary and preventive measures is heavily influenced by an individual’s knowledge, attitude, and practices (KAP) (Ajilore et al., 2017; Zhong et al., 2020). A KAP study is carried out with a specific population to gather information about their ideas, thinking, and execution to a specific object (WHO, 2008). Several studies revealed that the previous SARS outbreak’s prevention was somewhat related to people’s knowledge and attitude toward infectious diseases (Tao, 2003; Person et al., 2004). Furthermore, there is a scarcity of information on Rohingya refugees’ perspectives on the COVID-19 pandemic situation in Bangladesh. Given the foregoing, we feel it necessary to assess the Rohingya population’s COVID-19-related knowledge, attitude, and practices during the pandemic. As a result, the current study aimed to evaluate the KAP of the Rohingya people in Bangladesh during the early stages of COVID-19 severity in Bangladesh. To the best of our knowledge, this is the first study to assess the KAP of Rohingya refugees in regards to COVID-19.

## Materials and methods

### Study design and participant selection

A cross-sectional study was conducted to assess the knowledge, attitude, and practices of Rohingya people in eight refugee camps of Ukhiya and Teknaf in Cox’s Bazar between July and September 2020. The only inclusion criterion was not being infected with COVID-19, and the only exclusion criterion was being under the age of 21.

### Sample size detection

The infinite population formula was used to calculate sample size [S = (Z)^2^ × P × (1-P) ÷ (M)^2^]. A 95% confidence level was used to calculate the Z-value (1.96). The population proportion (P) and margin of error (M) were calculated at the 50% (0.50) and 5% (0.05) levels, respectively. We increased the S from the calculated 385 to 400 Rohingya refugees, for an anticipated level of precision of 4%.

### Study tools and data collection

For convenience, the KAP questionnaire was prepared in two versions (English and Bengali). The survey was carried out with the assistance of 15 trained interviewers. After explaining the survey’s purpose and design, the interviewers approached the respondents and conducted a face-to-face interview to complete the questionnaire. The first part of the questionnaire was used to collect demographic information, and the second part was used to evaluate respondents’ KAP regarding COVID-19. To minimize the possibility of choosing the desired and correct answer by chance, three answering options were provided. The answering options in the knowledge section were a) true, b) false, and c) don’t know, whereas in the attitude section the options were a) agree, b) disagree, and c) not sure) and in the COVID-19 practices section the options were a) yes, b) no, and c) sometimes). Each correct and desired response was worth one point, while the remaining responses received no point or zero. The score ranges for the knowledge, attitude, and practices sections were 0-10, 0-5, and 0-5, respectively. After then the scores were converted to a scale of 0 to 100. A score of more than 60% was considered good, while a score of less than 60% was considered poor. The mean and standard deviation of the KAP scores are presented within the dataset. Furthermore, for ease of interpretation, response scores were converted into percentages.

### Statistical analysis

Before importing to the Statistical Package for Social Sciences (SPSS) software (version 20.0), all data were entered on a master Microsoft Excel spread sheet. To assess the differences in mean KAP scores between demographic variables, a paired sample t-test and ANOVA were used. A p-value of less than 0.05 was considered significant.

### Ethics statement

The institutional review committee of Cox’s Bazar Medical College and Hospital checked the design and approved the study. The participants’ implied consent to participate in the study was based on their verbal acceptance of the survey and their responses to the interviewers’ queries.

## Results

### Demographic characteristics

The study enrolled 400 people, with 187 (46.8 %) males and 213 (53.3%) females. Three hundred sixty-three (90.8%) of those polled had lived in the camp for more than a year. The majority of respondents (30.8%) were between the ages of 51 and 60, with those between the ages of 41 and 50 coming in second (29.3%). Two hundred sixty-two (65.5%) respondents had no formal education; however, 53 (13.3%), 55 (13.8%), and 30 (7.5%) had less than 2, 2 to 5, and more than 5 years of education, respectively. More than half (59.8%) were married and 30.5% were single (**Table 1**).

**Table 1.** Demographic characteristics of survey participants (n=400)

| Variables/Characteristics | n (%) |
| --- | --- |
| Gender |  |
| Male | 187 (46.8) |
| Female | 213 (53.3) |
| Age (year) |  |
| 21-30 | 70 (17.5) |
| 31-40 | 90 (22.5) |
| 41-50 | 117 (29.3) |
| 51-60 | 123 (30.8) |
| Time spent in the camp |  |
| < 1 year | 37 (9.3) |
| > 1 year | 363 (90.8) |
| Marital Status |  |
| Married | 239 (59.8) |
| Unmarried | 122 (30.5) |
| Others (Divorced, widow, etc.) | 39 (9.8) |
| Years of education received |  |
| No education | 262 (65.5) |
| < 2 years | 53 (13.3) |
| 2-5 years | 55 (13.8) |
| > 5 years | 30 (7.5) |

### COVID-19 knowledge

The majority of participants (307, or 76.8%) stated that they were aware that COVID-19 is caused by a virus. Three hundred forty-nine (87.3%) of those interviewed were familiar with the major symptoms of COVID-19 (fever, cough, and fatigue). The level of knowledge about COVID-19 transmitting through foods and COVID-19 being unable to penetrate cloth masks was low among study participants; only 156 (39.0%) and 137 (34.3%), respectively, knew that COVID-19 transmits through foods and is unable to penetrate cloth masks. COVID-19 could be spread by respiratory droplets, was known to 269 (67.3%) of respondents. More than half (208, or 52%) were unaware that people with heart disease, diabetes, and high blood pressure are more likely to be infected with COVID-19. A remarkable number of people (294, or 73.5%) agreed that isolating and treating infected people are effective ways of limiting virus spread. Almost two-thirds (258, or 64.5%) were aware that there is still no permanent cure for COVID-19. On the other hand, 173 (43.3%) of participants were informed that COVID-19 is the same as the flu virus and that not all COVID-19 infected people will experience severe symptoms (**Table 2**).

**Table 2.** Responses of participants regarding COVID-19 knowledge

| Questions | Correct answer,<br>n (%) | Wrong answer,<br>n (%) |
| --- | --- | --- |
| 1. COVID-19 is caused by virus | 307 (76.8) | 93 (23.3) |
| 2. Fever, cough, fatigue are major symptoms of COVID-19 | 349 (87.3) | 51 (12.8) |
| 3. COVID-19 can transmit through foods | 156 (39.0) | 244 (61.0) |
| 4. COVID-19 can spread through respiratory droplets | 269 (67.3) | 131 (32.8) |
| 5. People with heart, diabetes and high blood pressure are more likely to be infected by COVID-19 | 192 (48.0) | 208 (52.0) |
| 6. COVID-19 cannot penetrate the cloth masks | 137 (34.3) | 263 (65.8) |
| 7. Isolation and treatment of infected people are effective methods of limiting virus spread | 294 (73.5) | 106 (26.5) |
| 8. COVID-19 is same as flu virus | 173 (43.3) | 227 (56.8) |
| 9. Currently there is no effective cure for COVID-19 | 258 (64.5) | 142 (35.5) |
| 10. Not everyone infected with COVID-19 will develop severe symptoms | 173 (43.3) | 227 (56.8) |
| Score, Mean (SD) | 5.8 (1.8) |  |

### COVID-19 attitude and practices

**Table 3** shows the attitude of Rohingya people regarding COVID-19. A good number of respondents (268, or 67%) agreed that wearing mask is very important during this pandemic time. Hand washing and the use of sanitizer were the most frequently reported methods of avoiding COVID-19 virus infection, and 238 (59.5%) of respondents agreed on this. Lockdown is required to control the spread of COVID-19, according to nearly 40% of respondents (159, or 39.8%). However, nearly 90% (342, or 85.5%) of them were unsure about staying at home while suffering from COVID-19 symptoms. 167 (41.8%) respondents were concerned that COVID-19 would spread throughout the camp settings.

**Table 3.** Participants’ attitude responses towards COVID-19.

| Questions | Agree,<br>n (%) | Disagree,<br>n (%) | Not sure,<br>n (%) |
| --- | --- | --- | --- |
| 1. Imposing lockdown is necessary to control the spread of COVID-19 | 159 (39.8) | 82 (20.5) | 159 (39.8) |
| 2. Wearing mask is very important in this pandemic time | 268 (67.0) | 53 (13.3) | 79 (19.8) |
| 3. I am worried that COVID-19 will spread throughout the camp settings | 167 (41.8) | 65 (16.3) | 168 (42.0) |
| 4. Regular hand washing and the use of sanitizer can safeguard an individual during COVID-19 | 238 (59.5) | 34 (8.5) | 128 (32.0) |
| 5. If you have the symptoms, you should stay at home | 58 (14.5) | 112 (28.0) | 230 (57.5) |
| Score, Mean (SD) | 2.2 (1.0) |  |  |

**Table 4** displays the COVID-19 practice of the interviewed participants. When asked if they frequently went in a crowded place during COVID-19, more than half of respondents (227, or 56.8%) said they did. When it came to shaking hands with outsiders, about 90% of the refugees (352, or 88%) did not appear to be very careful. Compared to this, a large number of people (128, or 32%) wear masks when they go outside. Nearly 60% (221, or 55.3%) of respondents said they occasionally washed their hands after coming in from outside, while 31% (n=124) said they never did. Below 40% (135, or 33.8%) of participants said that they covered their face while coughing or sneezing

**Table 4.**
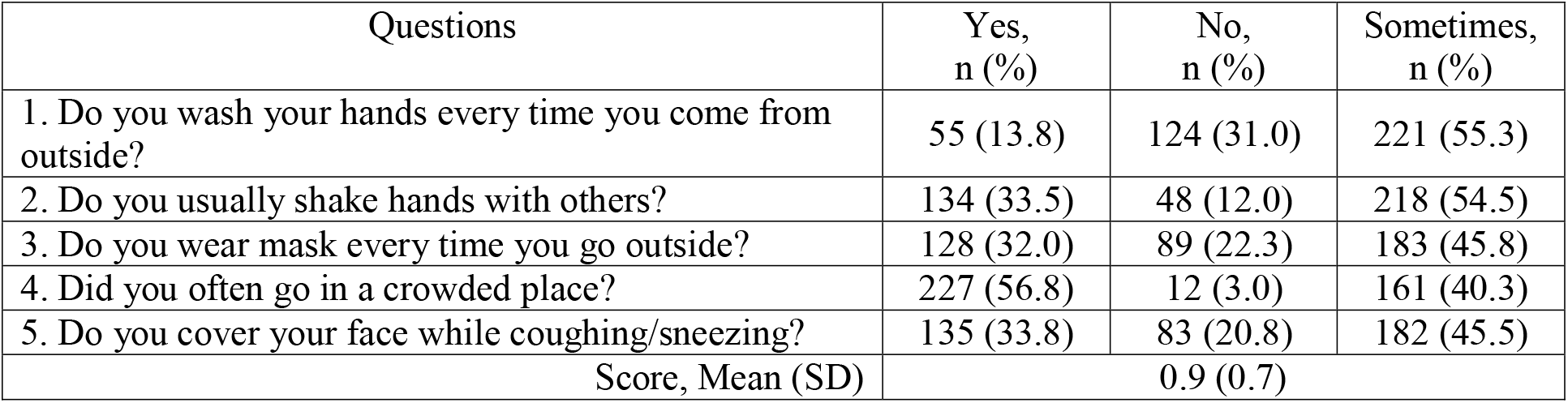
Responses of participants regarding COVID-19 related practices.

### Sources of COVID-19 information

The NGOs working inside the refugee camps played a critical role in disseminating information about COVID-19 to the people. According to **Figure 1**, the majority of those who took part in this study (172, or 43%) received the COVID-19 information from the NGOs. As a source of receiving information about the pandemic, social media came in second place. About 27% (07, or 26.8%) people obtained relevant information from different social media. Rest of the participants got the message from television, newspapers, radios, poster, banners, and leaflets.

**Figure 1.**
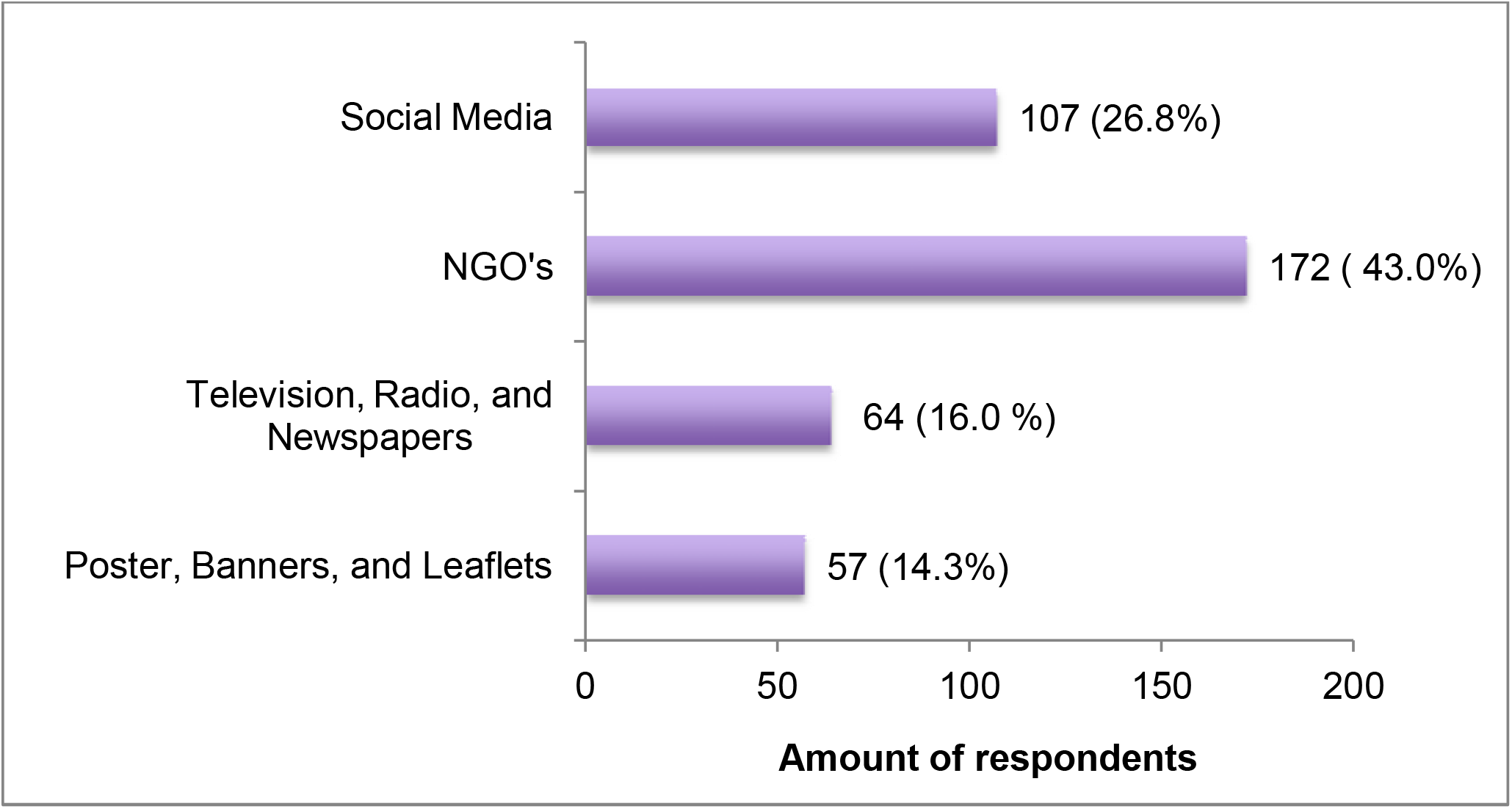
Sources of information about COVID-19.

### Association of KAP scores with demographic variables

There was a significant difference in knowledge between male and female respondents. Male participants scored significantly higher on their COVID-19 knowledge. In the case of COVID-19 attitude and practices, no significant difference was observed in them. There was no significant difference in KAP scores based on age, length of stay, or marital status. However, respondents aged 21-30, on the other hand, had higher KAP scores. On the other hand, those with more than 5 years of education, scored significantly higher in COVID-19 knowledge, but the scores for attitude and practices did not differ significantly based on education.

## Discussion

To the best of the author’s knowledge, this is the first ever study that assesses the KAP of Rohingya refugee people regarding COVID-19. We, therefore, attempt to discuss and compare our findings with those of other refugee studies as well as other generic Rohingya KAP studies. A KAP survey can be beneficial if conducted in the early phase of a situation (WHO, 2008). This KAP study began in the second month after the first COVID-19 case was identified (Jubayer et al., 2021) in the Rohingya camps in Cox’s Bazar, Bangladesh. Improved sanitation facilities, as well as very good knowledge, attitude, and, most importantly, consistent and accurate practice of hygiene and sanitation methods, are critical for disease outbreak prevention and control (Nahimana et al., 2017).

In the current study, we observed that the Rohingya participants had a very poor attitude (2.2 ± 1.0) and practices (0.9 ± 0.7) toward COVID-19, whereas their knowledge (5.8 ±1.8) was found to be slightly improved when compared to these two. A study was conducted recently on Rohingya people in the Cox’s Bazar camps regarding water, sanitation, and hygiene (WASH), and similar results were found in the overall knowledge scores (Hsan et al., 2019). In the knowledge section, we found in five questions about half of the respondents failed to provide correct answers. The questions concerned the transmission of COVID-19 through foods, the effectiveness of cloth masks, comorbidities, the severity of developed symptoms, and whether COVID-19 is similar to the flu virus. Similarly, in another study, respondents were unable to correctly answer transmission-related questions (Hamadneh et al., 2021). Respondents with some educational qualifications appeared to perform well in these types of COVID-19 questions (Alnasser et al., 2021; Al-Hussami et al., 2021; Banik et al., 2021; Huynh et al., 2020; Jadoo et al., 2020; Yue et al., 2021). The majority of the participants in this study (65.5%) were uneducated. Another study (Haque et al., 2020) on the Rohingya people of Bangladesh discovered a higher rate of illiteracy among respondents (79.6%). Haque et al., 2020 also stated that a reasonable impact on knowledge and behavior necessitates an educational background. There is a significant difference in the knowledge scores based on education (**Table 5).** A lack of information during the period of rapid rise may be another possible cause (Hamadneh et al., 2021).

**Table 5.** Comparison of KAP scores and demographic variables.

| Variables | Knowledge score<sup>a</sup> | p-value | Attitude score<sup>a</sup> | p-value | Practices score<sup>a</sup> | p-value |
| --- | --- | --- | --- | --- | --- | --- |
| Gender |  |  |  |  |  |  |
| Male | 6.1 (1.8) | 0.041<sup>b</sup> | 2.4 (1.2) | 0.071<sup>b</sup> | 1.1 (0.8) | 0.269<sup>b</sup> |
| Female | 5.4 (2.0) |  | 1.9 (1.0) |  | 0.8 (0.7) |  |
| Age (year) |  |  |  |  |  |  |
| 21-30 | 6.7 (1.9) | 0.126<sup>c</sup> | 2.9 (1.2) | 0.206<sup>c</sup> | 1.1 (0.6) | 0.730<sup>c</sup> |
| 31-40 | 6.5 (2.0) |  | 2.8 (1.2) |  | 1.0 (0.7) |  |
| 41-50 | 5.7 (1.8) |  | 2.0 (1.1) |  | 1.1 (1.0) |  |
| 51-60 | 4.8 (1.8) |  | 1.6 (0.8) |  | 0.7 (0.5) |  |
| Time spent in the camp |  |  |  |  |  |  |
| < 1 year | 5.2 (1.3) | 0.143<sup>b</sup> | 2.4 (1.1) | 0.692<sup>b</sup> | 0.9 (0.5) | 0.993<sup>b</sup> |
| > 1 year | 5.8 (1.9) |  | 2.2 (1.0) |  | 0.9 (0.7) |  |
| Marital Status |  |  |  |  |  |  |
| Married | 5.8 (1.9) | 0.923<sup>c</sup> | 2.2 (1.1) | 0.964<sup>c</sup> | 1.0 (0.7) | 0.862<sup>c</sup> |
| Unmarried | 5.8 (1.7) |  | 2.2 (0.9) |  | 0.8 (0.6) |  |
| Others (Divorced, widow, etc.) | 5.5 (1.8) |  | 2.4 (0.9) |  | 0.8 (0.5) |  |
| Years of education received |  |  |  |  |  |  |
| No education | 5.0 (1.9) | 0.019<sup>c</sup> | 1.7 (1.0) | 0.073<sup>c</sup> | 0.6 (0.6) | 0.206<sup>c</sup> |
| < 2 years | 7.2 (1.7) |  | 3.1 (1.1) |  | 1.4 (0.8) |  |
| 2-5 years | 7.1 (1.9) |  | 3.2 (1.0) |  | 1.5 (0.9) |  |
| > 5 years | 7.4 (1.8) |  | 3.3 (0.9) |  | 1.7 (0.8) |  |

<sup>a</sup> Data are presented as Mean (SD)
<sup>b</sup> Paired *t*-test; $p < 0.05$ considered significant
<sup>c</sup> ANOVA; $p < 0.05$ considered significant

According to our findings, the overall attitude of the Rohingya respondents toward COVID-19 was unsatisfactory. They exhibited a comparatively positive attitude toward the use of masks as well as hand washing and sanitizing issues. But when come to practice, we found a very little number of people (<20%) are not used to it. The hygiene behavior of the Rohingya participants in this study contradicts the findings of the study by Lopez-Pena et al., 2020. Hsan et al., 2019, on the other hand, stated that more than 50% of the Rohingya refugees engaged in unsafe hygiene practices. Haque et al., 2020 showed that the literacy rate and family size have no effect on the hygiene behaviour of the Rohingya people. Furthermore, finding a relationship in such a matter is irrelevant because the entire community is in desperate need of relief goods (Haque et al., 2020). There are scarcities of hygiene and sanitation items inside the camps. Notably, about 88% of Rohingya people depend on external aid from UN agencies and other non-governmental organisations to meet their needs of daily living (WFP, 2020). This could be one of the reasons why people are unable to practice proper hygiene and sanitation. However, the people do not want a lockdown in the camps, as approximately 60% of those polled responded negatively to this question. Regrettably, they do not believe that they should remain at home during this time. About two-thirds of them appeared unconcerned about spreading the disease within the camps. This attitude of them also reflects in their practices. More than 80% of people said they usually shake hands with others and go to crowded places. These kind of risky behaviors of the Rohingya people were also reported by Lopez-Pena et al., 2020. But, this is the polar opposite of the Syrian refugee mothers. They demonstrated a very positive attitude and practice in social distance and large gatherings (Hamadneh et al., 2021).

According to a recent study by Lopez-Pena et al., 2020, NGOs are trusted sources of information among Cox’s Bazar refugees. In the current study, approximately half of the participants (43%) said they learned about COVID-19 from NGOs working inside the camps. The second highest number of respondents (26.8%) in this present study obtained relevant information from various social media. However, according to one study, among Syrian refugee mothers, various social media platforms (Facebook, Whatsapp, etc.) were the most popular source of COVID-19 information (Hamadneh et al., 2021). In the refugee camps of Bangladesh, more than 100 NGOs are functioning (ISCG, 2020). The NGOs’ active participation in the COVID-19 may make them the primary source of information for the camp residents. On the other hand, the findings support the provision of social and print media within the camps for disseminating health information (Le et al., 2020).

The entire refugee accommodation area in Cox’s Bazar possesses the characteristics that make it likely to become an epicentre of infection spread. Current camp conditions include lack of social distancing, food insecurity, and scarcity of knowledge and awareness about COVID-19. A lack of personal protective equipment for health workers at other health facilities at Ukhiya, Teknaf and other camp health facilities creates a barrier to even primary medical consultation (Truelove et al., 2020). These situations may impact the knowledge, attitude, and practices of the Rohingya people regarding COVID-19.

## Data Availability

Data are available upon reasonable request

## Limitations and future directions

Because it measured their self-reported practices, this study is prone to respondent bias and may not reflect actual practice. Furthermore, we did not test the questionnaire’s validity and reliability. As for future directions, a strong training and behavior change communication program may help to improve the situation and prevent disease outbreaks. A follow-up KAP study with two groups of refugees (trained and untrained) could be conducted to improve training content and quality.

## Acknowledgements

We thank the interviewer for their kind assistance and time. We also appreciate the Rohingya people’s participation in this research.

## Funding information

This work received no funding.

## Data availability statement

Data are available upon reasonable request.

## Declaration of interest

The authors declare no conflicts of interest.

## Notes

### Competing Interest Statement

The authors have declared no competing interest.

### Clinical Trial

Not applicable

### Funding Statement

No funding received

### Author Declarations

The institutional review committee of Coxs Bazar Medical College and Hospital checked the design and approved the study. The participants implied consent to participate in the study was based on their verbal acceptance of the survey and their responses to the interviewers queries

